# Pilot Clinical Validation of a Machine Learning Platform for Noninvasive Smartphone-Based Assessment of Corneal Epithelial Integrity

**DOI:** 10.1101/2023.08.29.23293788

**Authors:** Andrew Y. Zhang, Jayanth S. Pratap, James R. Young, Joshua Lui, Kian Attari, Arnav A. Srivastava, Erica R. Wu, Aracely D. Moreno, Annie Miall, Jay M. Iyer, Sreekar Mantena, Jay Chandra, Vineet P. Joshi, Deborah S. Jacobs

## Abstract

**Purpose:** Fluorescein staining (FS) is a standard method of assessing corneal epithelium (CE) integrity. However, the equipment and personnel required for FS may be unavailable in low-resource environments. We developed and validated a low-cost, noninvasive, and quantitative CE evaluation pipeline using a custom smartphone attachment and convolutional neural networks (CNNs).

**Methods:** A 3D-printed smartphone attachment and placido disk illumination module was attached to a OnePlus 7 Pro smartphone. 26 smartphone-acquired images were obtained from 15 subjects, comprising a dataset including healthy eyes and corneal epitheliopathies of Oxford grade I-V. A classifier CNN was trained on 8 subjects (23,173 image patches) to identify areas of suspected epithelial disruption, and validated on 7 subjects (10,883 image patches). The fraction of disrupted corneal surface area (FDSA) was computed for each subject from the model output. Results were compared with FS slit lamp photos which were independently graded by two clinicians using the Oxford scheme.

**Results:** FDSA showed promise as a non-invasive marker of CE integrity, with mean FDSA in the Oxford >II cohort being higher than the Oxford ≤II cohort (*p = 0.04* and *p = 0.09* using Oxford scores from each clinician, respectively). Additionally, areas of CE disruption identified by our smartphone-based technique showed qualitative concordance with those revealed by FS.

**Conclusions:** Our technique for smartphone-based CE imaging and automated analysis is a promising low-cost, noninvasive method to quantitatively evaluate the CE.

**Translational Relevance:** This tool can be used to evaluate ocular surface disease in low-resource regions.

## INTRODUCTION

The cornea has a surface epithelium (CE) that creates a barrier to the outside environment and is critical to the eye’s refractive power and clarity^1^. The epithelium may be disrupted by trauma, infection, exposure, dry eye disease, systemic disease with ocular surface involvement, and topical medications and their preservatives. CE breakdown is typically painful and leaves the eye susceptible to invasion by pathogens.

Identifying and treating corneal epithelium breakdown is critical to ocular health and preservation of vision. Currently, corneal epithelium breakdown is diagnosed with fluorescein staining (FS). Fluorescein dye applied to the cornea adheres to exposed basement membrane and corneal stroma and is visualized using cobalt blue light and a slit lamp biomicroscope^2,3^. In low-resource regions, many of the health facilities that bear the brunt of the medical burden do not have access to specialized equipment or trained medical technicians. This problem is magnified by imbalances in healthcare coverage: in India, for instance, 60% of all medical personnel reside in urban centers despite these centers only containing 30% of the country’s population^4^. In addition, FS is generally assessed by visual inspection and even with standard grading systems may be ill-suited for longitudinal evaluation, where gradual changes may not always be visually apparent^5^. Finally, the grading process has been found to be inconsistent between different clinicians^6^. The management of corneal epitheliopathies can thus benefit from a portable, low-cost, and more quantitative means of evaluating the corneal surface.

In recent years, smartphone-based innovations have shown potential to expand access to ophthalmological screenings due to their inexpensive and scalable nature. For instance, a low-cost, point-of-care tool for evaluating corneal endothelial health has demonstrated comparable accuracy to “gold standard” specular microscopy, which costs tens of thousands of dollars^7^. Another smartphone-based device uses an illuminated 3D-printed attachment and an image analysis pipeline to diagnose keratoconus^8^. With regards to fluorescein examination, however, there does not yet exist a tool that is standalone, low-cost, quantitative, and noninvasive. Although computer vision algorithms have been developed to quantify FS images, they still require high-resolution digital image capture of the eye, which is typically performed by attaching a camera to a slit-lamp microscope^3,9^. Moreover, the use of fluorescein dye poses an additional resource requirement as well as the risk of patient discomfort.

To address this need, we developed a smartphone-based CE evaluation tool ideal for remote and low-resource settings. We designed modular adaptors that project an illuminated pattern onto the corneal surface, enabling the identification of intact and disrupted corneal surface regions based on their distinct reflective properties. We also trained convolutional neural networks (CNNs) to process images and highlight regions with suspected corneal surface disruption. The hardware attachments can be readily and cheaply produced using 3D-printed components and LEDs, and can be customized to accommodate a variety of phone dimensions.

Our novel platform enables rapid, point-of-care evaluation of corneal epithelial integrity. Unlike previous methods, our system does not require the use of fluorescein dye, thereby eliminating one of the many barriers of corneal imaging in low-resource communities. Furthermore, our hardware attachment is capable of being manufactured at less than $10 apiece, compared with a slit lamp which can cost thousands of dollars. In contrast to the current standard, our system is portable and image analysis does not require trained professionals, allowing for expanded access to CE evaluation for follow-up treatment or prophylactic care.

## METHODS

The corneal surface yields evidence of any surface disruptions via distortions in the reflection of a projected image. The projection of a high-density illumination pattern (HDIP) onto a healthy corneal surface yields a reflection which perfectly recreates the pattern, whereas any surface disruptions (for example, due to infection or dry eye) will result in imperfections in the reflection. Disrupted regions of arbitrary size will be visible in the reflection so long as the HDIP is of sufficiently high spatial frequency. By capturing the reflection with a smartphone camera and processing the image with CNNs, we identify and quantify regions of suspected epithelial disruption. In this section, we present elements of our hardware and software design and their motivating factors.

### Hardware

The smartphone attachment is composed of an attachment body and an HDIP module. Current prototypes are 3D-printed in polylactic acid (PLA), though our design is amenable to other manufacturing methods such as injection molding. The attachment body is a two-piece plastic clamp which has a snap-on interface for the HDIP module as well as built-in grips for holding the device in a vertical or horizontal orientation (**Fig. 1a-b**). Although in principle the attachment body can be made adjustable such that it can accommodate multiple smartphone models, during testing we found that a model-specific design with no moving parts was easier to use and more feasible given the substantially lower manufacturing costs. In practice, different attachment body designs to accommodate different smartphone models can be readily and cheaply produced. In the prototype shown here, the attachment body is designed for a OnePlus 7 Pro smartphone (OnePlus Technology, Shenzhen, China).

**Figure 1.**
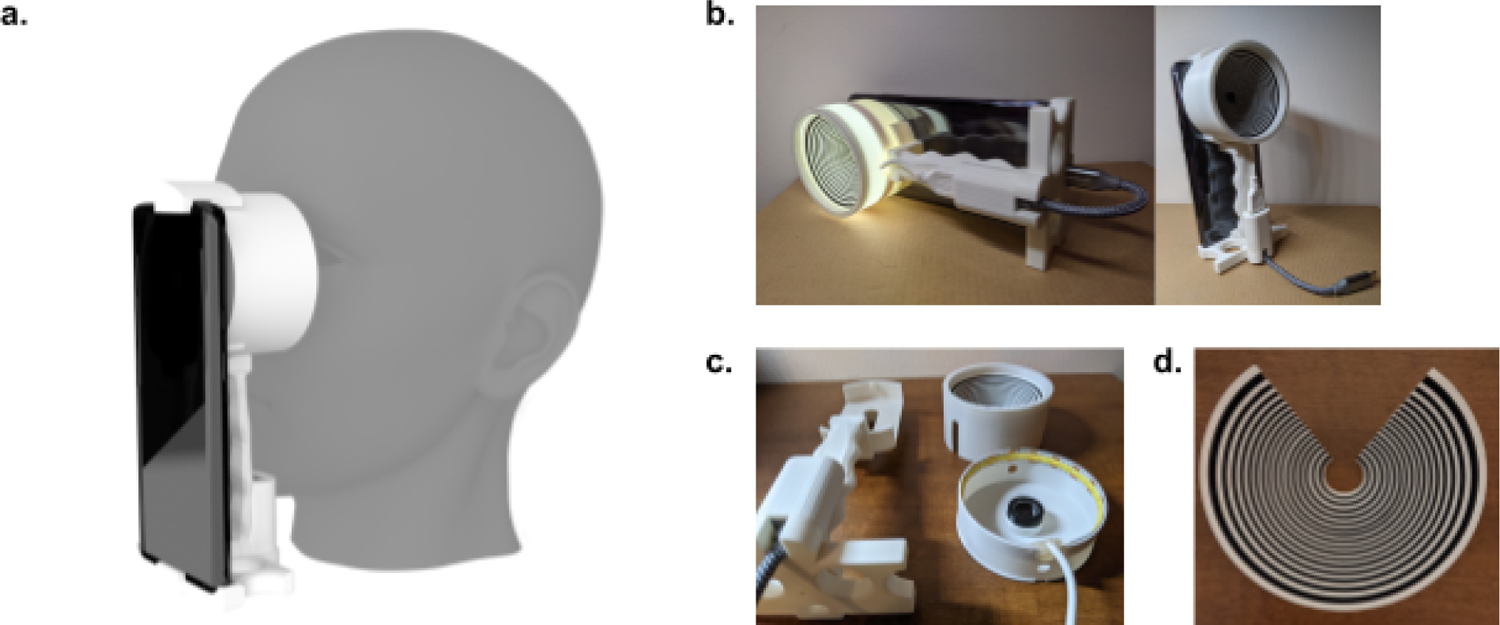
Smartphone attachment. a) 3D render of device with attached OnePlus 7 Pro smartphone. b) Photos of the device with illumination switched on or off. c) Disassembled components of device. d) Close-up of placido disk insert.

The HDIP module projects a pattern onto the corneal surface and enables the reflection to be captured by the smartphone camera. It consists of a base, lens insert, and diffuser (**Fig. 1c-d**). A chip-on-board (COB) flexible LED strip (Dephen, Shenzhen, China) is mounted in a circular ring on the inner wall of the base, and is attached to a USB A-to-C adapter (JSAUX, Shenzhen, China), which can draw power from either a 5V power bank or directly from the attached smartphone. The base also contains a mounting point for a convex lens with diameter 12.5 mm and focal length 35 mm (Part #L5865, Surplus Shed, Fleetwood, PA). This lens decreases the minimum focus distance of the smartphone camera to enable it to focus on the corneal surface. The diffuser is a double-walled conical shell which serves to diffuse the light from the LED strip to evenly illuminate the cornea, and it holds an interchangeable insert that creates the HDIP. Diffusers with different heights may be produced to allow for variation in the depth of the subject’s ocular orbit.

Although many patterns for the HDIP insert are possible, we adopted a placido disk due to it being radially symmetrical, and thus more amenable to downstream image processing algorithms (**Fig. 1d**). Moreover, the spatial frequency of a placido disk is readily adjustable by altering the thickness of the circular mires. In the prototype shown here, the insert is made from paper-backed vinyl (Craftables, Springfield, TN) from which alternating rings have been cut and removed from the upper vinyl layer. Other materials for the insert are possible, including printer paper or cardstock on which the placido pattern is printed, as long as the pattern can be projected with sufficient contrast between light and dark mires. The density of the placido mires determines the smallest disrupted region able to be resolved by this system.

Due to the geometry of the placido-cornea system, evenly-spaced mires on the placido insert will result in unevenly-spaced mires on the corneal reflection. In order to maximize the performance of the image processing algorithm, the spacing of mires on the reflection must be as uniform as possible. To solve this issue, we wrote a custom Python ray-tracing script which calculates the spacing of mires on the placido insert required to achieve a near-uniformly spaced reflection. Given the dimensions of the HDIP module and the desired mire density, the script computes the angles of the projected and reflected rays and generates an SVG image for printing or vinyl cutting (**Fig. S1**). In this script, the corneal surface is approximated as a spherical convex mirror with radius 7.8 mm^8^.

### Software Overview

The software consists of a graphical user interface (GUI) frontend and two CNN models on the backend. The GUI allows the user to upload an image or video and manually select the frame (if video input) and region of interest (ROI), as well as batch process images (**Fig. 2**). As a proof of concept, to assist with ROI selection, a regression CNN can optionally be invoked to automatically estimate the *(x, y)* coordinates of the center of the mires (**Fig. S2**). A patch-based CNN classifier is then used to identify disrupted regions in the ROI (**Fig. S3**). All software was developed in Python 3.8.8 with Tensorflow version 2.8.0 and OpenCV version 4.5.5.

**Figure 2.**
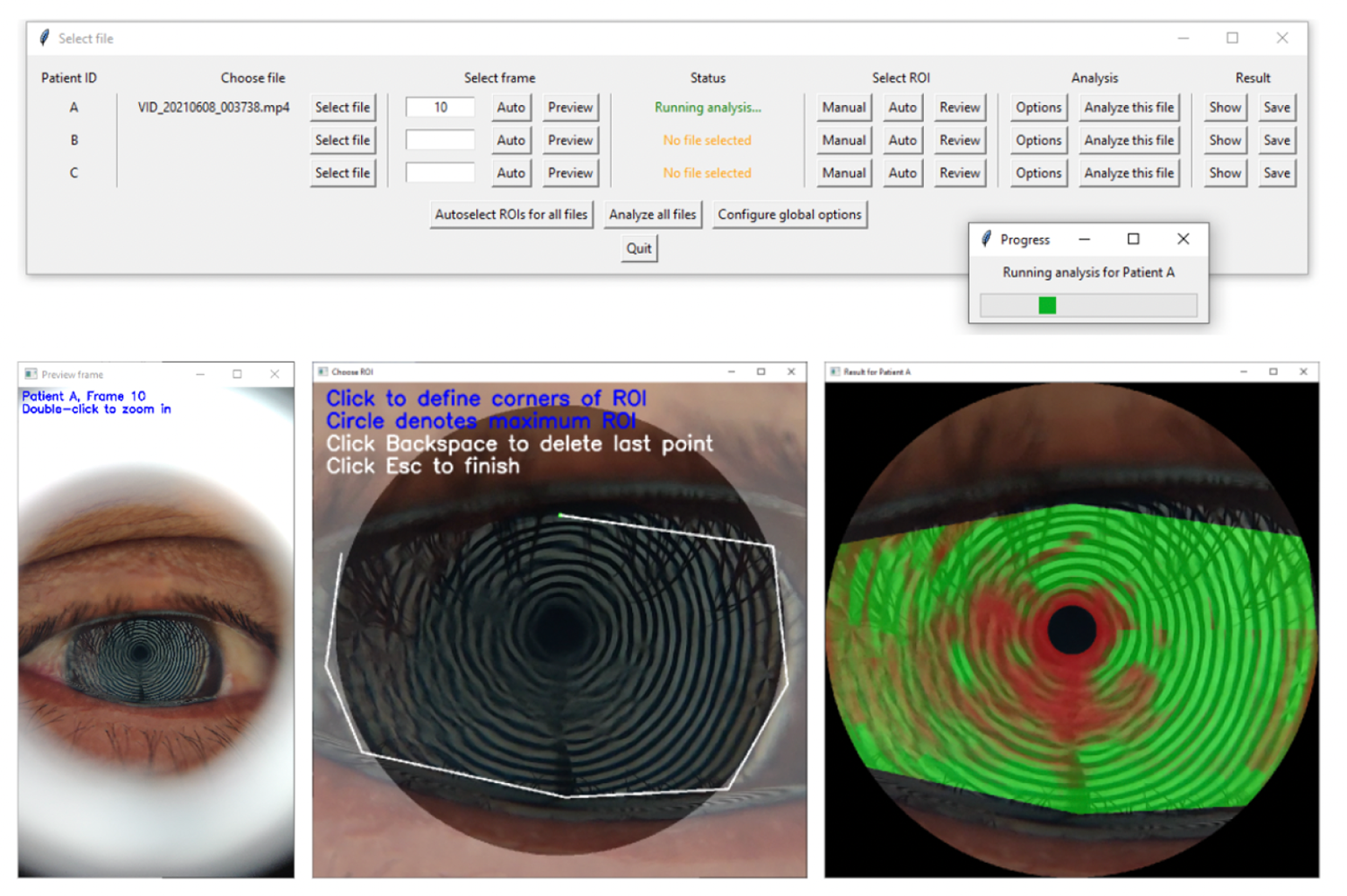
Graphical user interface for high-throughput screening of corneal subjects. Top row: main interface with progress bar. Bottom row, left to right: Frame preview, manual ROI selection interface, heatmap visualization of result.

### Data Acquisition

A pilot study was conducted at L.V. Prasad Eye Institute in Hyderabad, India. 15 subjects were imaged using the smartphone attachment in one or both eyes, forming a total dataset of 26 smartphone-captured images. All subjects were 18 years of age or older. Healthy patients as well as patients with corneal epitheliopathies of varying severity (Oxford I, II, III, IV, V) were represented within the data collected. This study was approved by the Institutional Review Board of the L.V. Prasad Eye Institute and informed consent was obtained from all subjects.

The hardware attachment was mounted on a OnePlus 7 Pro smartphone (OnePlus Technology, Shenzhen, China) and placed against the ocular orbit of each subject such that the optical axis of the camera was approximately in line with the optical axis of the pupil. The position of the device was adjusted until the reflection of the placido mires on the cornea came into focus. A short video at 4K resolution, 30 frames per second was captured, during which the subject was asked to blink. The process was repeated until a video with sufficient clarity was obtained. The video capture process took approximately 10 seconds per eye. A traditional fluorescein examination was also performed on the same eye.

### Regression Model Details

The exact center of the placido mires was manually annotated for 214 video frames from 6 subjects. Each image was first cropped to a centrally located 1000 × 1000 pixel region, then downsized to 500 × 500 pixels. Data augmentation was performed by shifting the initial location of the 1000 × 1000 pixel crop, which helps the model learn the location of the center regardless of its position within the input image. After augmentation, all images from one subject (336) were reserved for testing and the remaining images (2508) were used for training. Two neurons in the output layer were trained to predict the *x* and *y* coordinates of the center, respectively (**Fig. S2**).

### Classifier Model Details

In contrast to a typical semantic segmentation architecture such as a U-Net^10^, our patch-based classification approach allows the model to assign values between 0 and 1 to ambiguous regions. For each input image, a ground truth mask was manually drawn using GNU Image Manipulation Program (GIMP 2.10.22). Red and green were used to denote completely distorted and intact mire regions, respectively, while ambiguous regions were left unmarked (**Fig. S3**). During preprocessing, each frame was cropped to a 1000 × 1000 pixel square centered at the placido disk origin (either manually selected or estimated using the regression model above) and remapped to polar coordinate space while maintaining the same image dimensions. Every k = 20 pixels in the remapped image was sampled as a 3-channel patch of size 101 × 101 pixels, normalized, and mapped to the cropped and polar-remapped ground truth (**Fig. S3**). If the percentage of green pixels or the percentage of red pixels in the ground truth patch surpassed a predefined threshold of 80%, the patch was assigned to the “intact” or “disrupted” set, respectively (**Fig. S3**). The model was trained on this binary classification task, considering each patch independently of the others.

During inference, an image (after ROI masking) is preprocessed in the same manner as described above (**Fig. 3**). The single-neuron output layer yields a value between 0 and 1 for each patch, which is converted into an RGB value and assigned to the pixel at the patch center (where 1 corresponds to a pure green pixel, for example). Because every *k = 20* pixels is sampled as a patch (yielding 2500 total patches per 1000 × 1000 pixel image), the colors of areas between patch centers are linearly interpolated in order to generate a heatmap. The heatmap may be post-processed by masking out regions of the image which are not covered by the placido disk reflection. This heatmap is then overlaid on the original image to highlight predicted disrupted areas in red (**Fig. 3**).

**Figure 3.**
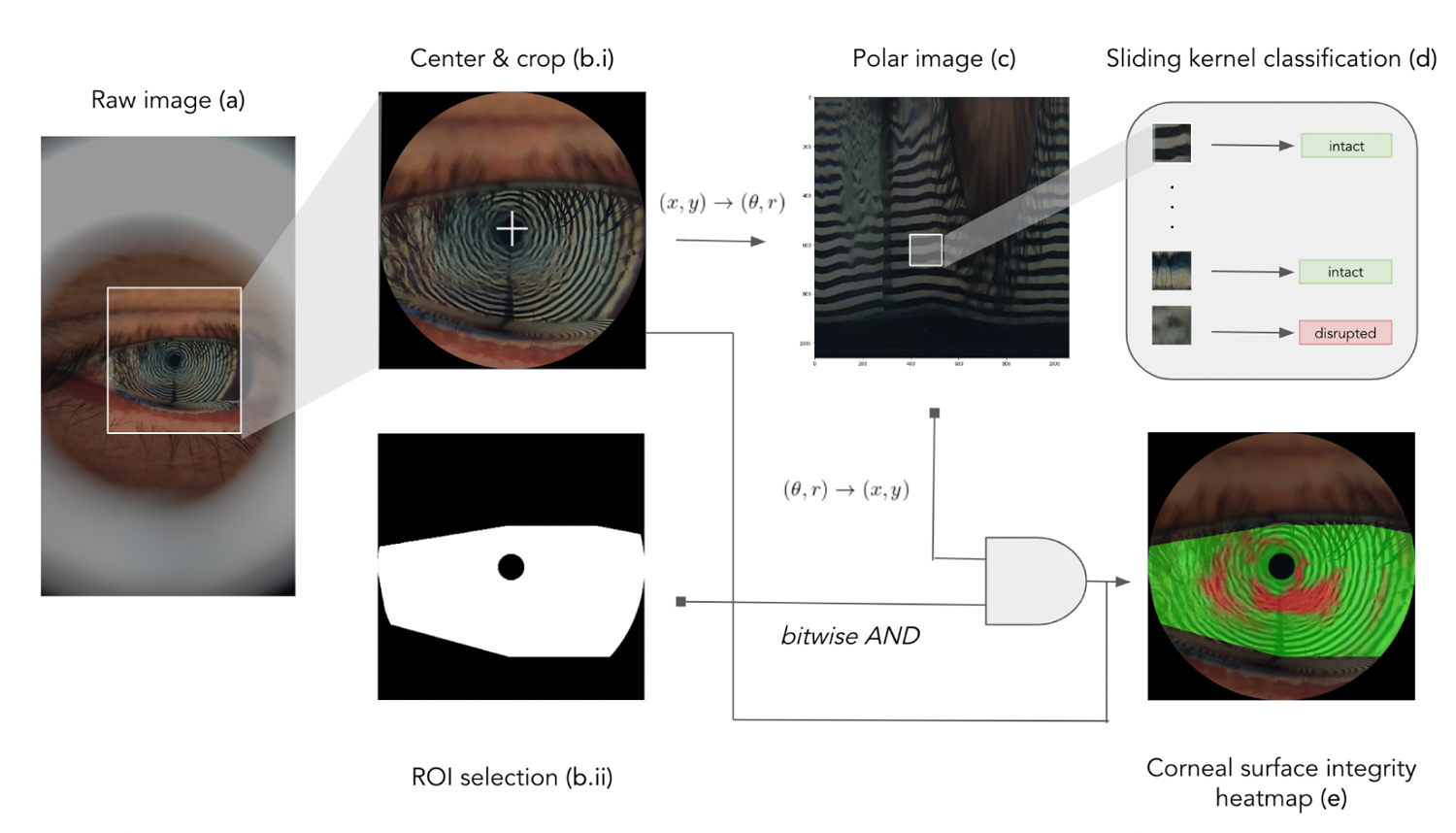
Image processing pipeline for epithelial defect identification from smartphone-acquired images. (a) Image capture of corneal surface; (b.i) automated center-finding algorithm with regression CNN or manual center annotation, followed by 1000×1000 circular crop; (b.ii) automated corneal segmentation *or* manual ROI selection; (c) transformation from Cartesian to polar coordinates; (d) patch-based inference with classification CNN; (e) postprocessing and masking to generate final heatmap visualization of surface integrity.

The classifier model was trained as follows. Each video was manually evaluated to determine the timestamps of any eye blinks present. The first frame with acceptable clarity was identified within the 1 second period following the blink, and if such a frame could not be found, the video was discarded. The pixel coordinates of the center of the placido disk reflection were manually annotated for each frame. In total, 26 frames from 26 videos representing 15 subjects were obtained for model training and evaluation. Finally, a stratified train-test split was performed manually such that 1) videos from the same subject belonged to the same dataset and 2) each dataset had an approximately equal distribution of intact and disrupted regions. In the training set, we allowed redundant frames (multiple frames representing the same eye), but in the testing set only one frame was kept per eye. This gave a total of 23,173 training patches from 8 subjects (17 frames) and 10,883 testing patches from 7 subjects (8 frames, not including one discarded frame from a duplicate eye). The proportion of intact patches was ∼77% in the training set and ∼81% in the testing set. Oxford grading^11^ was performed on the unpatched test set by two independent ophthalmologists who were not shown the output of the model.

### Statistical Analysis

For each eye, we computed the fraction disrupted surface area (FDSA), which is defined as the proportion of red pixels in the ROI-masked heatmap relative to the total number of nonzero pixels (where “red” is defined by an RGB value where *R > G*). The formula for FDSA is shown below, where *N* is the number of nonzero pixels, 𝟙 is an indicator function, and *R_i_*and *G_i_* represent the respective RGB values for the red and green color channels of pixel *i*.

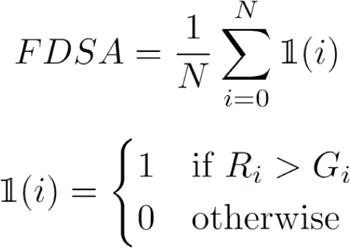

An unpaired one-tailed *t*-test was performed between the FDSA scores of Oxford ≤II and Oxford >II subjects, using the SciPy package (1.7.3) in Python (3.8.8). *P* values < 0.05 were considered statistically significant.

## RESULTS

The regression model was trained for 20 epochs, with a final MSE training loss of 6.09 px^2^ and testing loss of 574 px^2^. This indicates that, on average for this test set, the estimated center is about 24 pixels off from the true center (or 4.8% of the side length of the input image). Visually, the predictions are very close to the true center (**Fig. S2**). Inference for the regression model takes less than one second per image on an Intel Core i7-8706G CPU. Although the regression model was not invoked when testing the classifier model below, our results demonstrate that an automatic center-finding algorithm is feasible.

The classifier model was trained for 4 epochs, with a final accuracy of 90.87% across all patches of the test set. Inference for the classifier takes approximately 120 seconds per image on an Intel Core i7-8706G CPU. Visual inspection of the resulting heatmaps indicates good agreement with FS for subjects A, C, D, F, G with focal staining (**Fig. 4**). On the other hand, scattered or punctate staining patterns such as those found in subjects B and E are not well recapitulated by the heatmaps (**Fig. 4**). In particular, it appears that in the latter case the placido images either do not fully capture the diffuse nature of surface disruptions or introduce artifacts arising from poor focus or tear film residue. In nearly all cases, however, there is good agreement between the heatmaps and the disruption pattern present on the placido images. We note an exception to this for subject E, left eye, where the algorithm erroneously marks the darker pupillary region as disrupted. Model performance will likely improve with the inclusion of additional training data.

**Figure 4.**
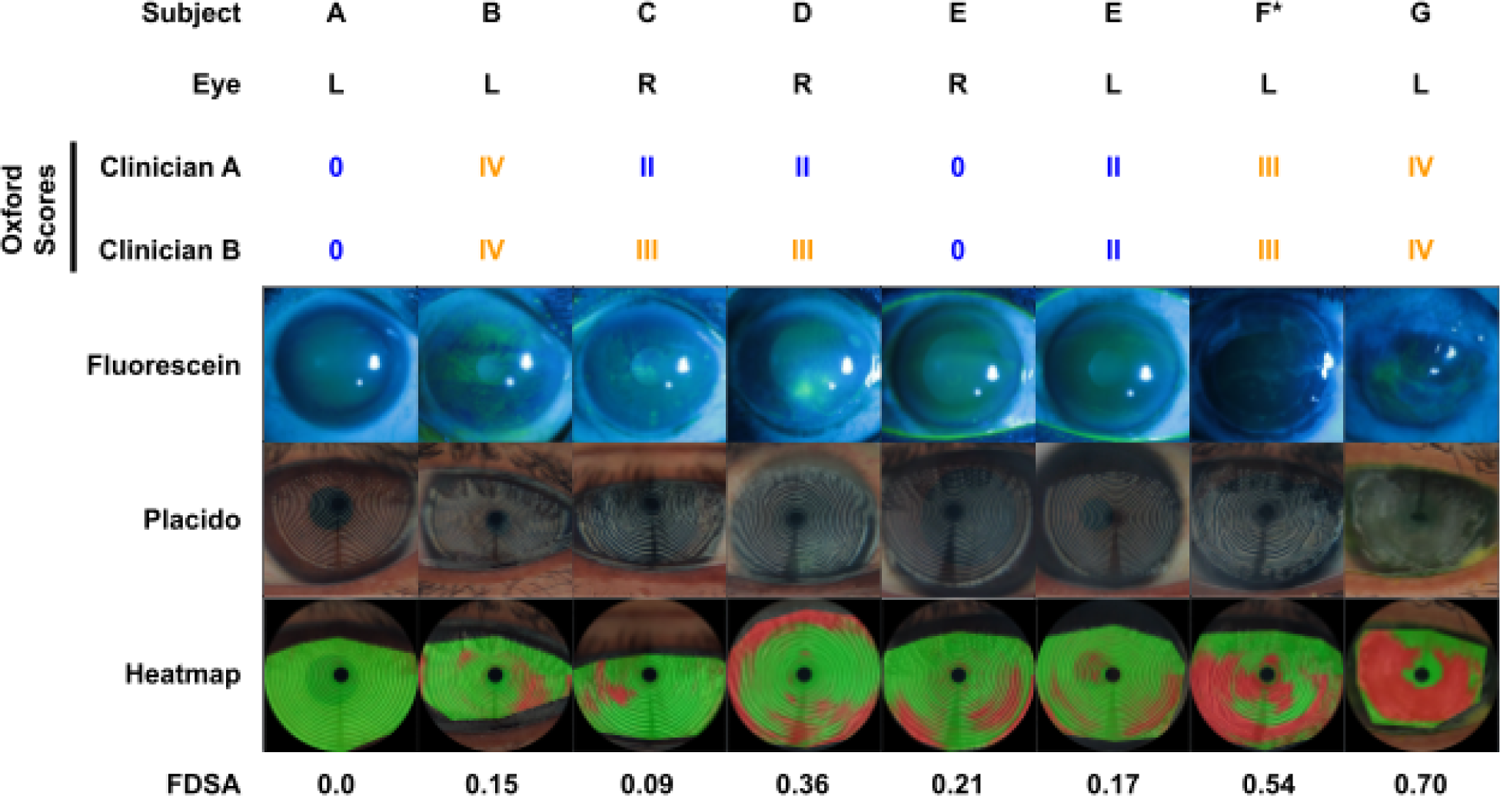
Epithelial defects revealed by fluorescein examination are visible under smartphone-based white-light imaging and are highlighted by the deep learning-based image processing pipeline. Image rows, top to bottom: Corneal epithelial defects visualized with fluorescein staining; placido disk images captured with handheld smartphone attachment; corneal epithelial defects computed by classifier network with manual ROI selection. Oxford scores were assigned independently by two ophthalmologists from the fluorescein images. Scores ≤II are shown in blue, scores >II are shown in orange. FDSA, fraction disrupted surface area (computed by algorithm). * indicates post-keratoplasty.

We compared average FDSA values between Oxford ≤II and Oxford >II subjects. Oxford scores were in agreement between the two clinicians except for subjects C and D, where clinician A assigned a score of II and clinician B assigned a score of III for each. Using clinician A’s Oxford scores, the average FDSA for the two classes was 0.166 and 0.463, respectively (one-tailed unpaired *t* test, *p = 0.04*). Using clinician B’s Oxford scores, the average FDSA for the two classes was 0.127 and 0.368, respectively (one-tailed unpaired *t* test, *p = 0.09*). Though the power of this analysis is limited by the small size of the labeled validation cohort, these results suggest that the FDSA statistic, calculated from the model output, is able to partially recapitulate the clinical insight from FS.

## DISCUSSION

In this study, we report the development of a novel and low-cost method of evaluating the health of the corneal epithelium. A 3D-printed smartphone attachment projects a placido disk pattern onto the corneal surface, and the reflection is captured by a smartphone camera. A companion software program automatically locates the reflected pattern in the resulting image and identifies regions of suspected corneal epithelial disruption. 15 subjects were imaged using the proposed system and, although quantitative results are limited due to the small sample size, we demonstrate the following proof-of-concept:

1. A reflected placido disk pattern reveals continuity of the corneal epithelium and is capable of highlighting irregular ocular surfaces.
2. A smartphone-based imaging system is capable of generating and capturing clinically useful images of the corneal epithelium.
3. A software pipeline can preprocess and automatically highlight regions of suspected damage in such images.

In general, the method performs better on conditions exhibiting focal FS, such as corneal ulcers, compared with those exhibiting scattered staining, such as superficial punctate keratitis. With additional training data and hardware refinement, accuracy of both the classification and regression models may be increased. This technology has the potential to simplify and standardize the diagnosis and monitoring of ocular surface disease.

The requirements of fluorescein dye and slit-lamp microscopy in the current standard of care for evaluating corneal surface integrity pose challenges in remote areas and resource-limited primary care settings, as well as in the care of subjects who are bedridden due to illness and who cannot travel to an eye clinic to be evaluated. In addition, the subjectivity of the fluorescein scoring procedure makes it difficult to track the progression of a corneal surface defect over time. Furthermore, ophthalmologists have historically had a higher level of patient-to-provider infections than other specialties, a risk exacerbated by COVID-19^12,13,14^. Using our proposed system, patients can be examined with minimal contact and without the need to administer fluorescein, potentially reducing disease transmission.

We now discuss current limitations of the proposed method, along with possible solutions. Firstly, the resolution of the placido mires is not sufficient for detection of smaller corneal surface disruptions, such as those found in superficial punctate keratitis; however, this resolution is sufficient for screening for large-scale defects, and could be increased by increasing the density of the placido mires. Secondly, there is an optical blind spot at the center of the placido disk, where the camera is located. However, this only covers around ∼1% of the placido reflection (∼12 mm^2^), and this limitation is inherent to the design of similarly designed corneal topographers used in clinics today^15^. If necessary, this could simply be resolved by capturing two corneal images at a slightly different angle and merging the result. Finally, a fundamental challenge when acquiring images is the need to ensure the reflection is in focus, which could be mitigated by integrating a customized autofocusing system in the image capture methodology.

Some additional challenges arise from our low-cost prototyping and manufacturing process. Firstly, because our placido insert is created from a two-dimensional sheet which is rolled into a cone, there are potential distortions and artifacts in the mire pattern if the insert is not properly seated in the diffuser. Secondly, as shown by our raytracing script (**Fig. S1b**), the placido disk reflection does not exist in one focal plane, and in fact occupies a convex surface. When focusing on the center of the cornea, this can cause reflected mires at the periphery of the placido pattern to appear slightly out of focus. These issues may be addressed by 1) printing the placido disk pattern onto the diffuser directly, and 2) experimenting with non-conical designs for the diffuser such that the virtual image of its projection forms a flat surface. These would require a modest increase in the cost of manufacture, but the total cost would still be far below that of traditional instrumentation.

With improvements in the hardware and additional data collection, additional software capabilities are planned for the near future. Firstly, additional image processing algorithms may be developed to enable automatic image capture by detecting blinks and verifying the image is in focus. The regression CNN presented here may be augmented with a segmentation model to enable automatic masking of non-mire regions. Given the morphology and extent of disrupted corneal regions, another deep learning model may be trained to provide both Oxford/NEI-equivalent scores^11,16^ as well as etiological diagnoses. For example, herpes simplex keratitis yields a distinctive dendritic ulcer^17^. These capabilities would allow this method to be more easily integrated within existing clinical workflows. Finally, we plan to convert the software into a mobile application to run on-device for rapid, point-of-care evaluation without the need for an internet connection.

The development of mobile health technologies is a part of a broader ongoing initiative to deploy more tractable, scalable, and cost-effective healthcare solutions in remote locations. Opportunities for using mobile health technologies have significantly improved within the past few years with smartphones becoming an increasingly ubiquitous technology. Additionally, the use of digital imaging techniques is spreading worldwide, bringing mobile imaging to health clinics in remote rural areas. The increasing quality and reliability of machine learning algorithms, along with the falling cost of hardware, paves the way for transformed health care paradigms. Our novel low-cost noninvasive corneal imaging system offers an innovative solution to eye care challenges posed in low-resource environments and has the potential to advance global health equity.

## FUNDING STATEMENT

Funding for this study was provided by the Seva Foundation.

## Data Availability

All data produced in the present study are available upon reasonable request to the authors.

## ACKNOWLEDGEMENTS

The authors thank the participants of this study for their time, thank the Seva Foundation for their support, and thank the advisory board of the Global Alliance for Medical Innovation (GAMI) for their thoughtful feedback.

## SUPPLEMENTARY MATERIAL

**Figure S1.**
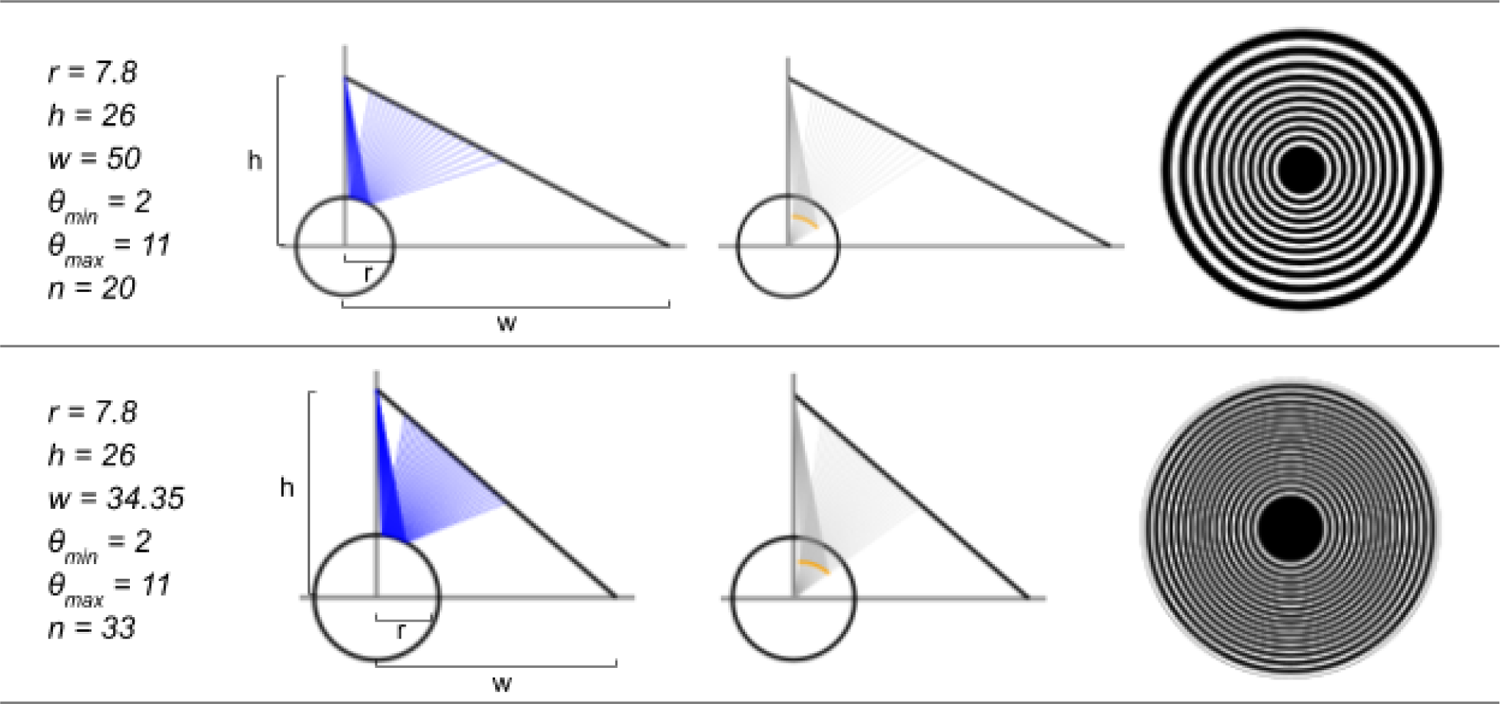
Design of white light illumination module, assisted by custom ray-tracing script. Two example placido disk designs are shown with their corresponding parameters. Diagrams show cross-section of optical system, with circle approximating the cornea and sloped line representing the conical wall of the diffuser. The position of the camera is approximated as the apex of the cone. The intersection points of the primary rays (blue) denote the approximately equidistant white-black mire transitions in the placido disk reflection on the cornea. The intersection points of the secondary rays (gray) represent the virtual image formed by the placido disk reflection. This virtual image appears as a convex surface (orange). The placido pattern is projected to 2D by the script (equivalent to flattening the conical surface) and shown in the rightmost column. Measurements: *r*, radius of circle; *h*, height of cone; *w*, radius of cone; *θ_min_*, angle of smallest ring from optical axis; *θ_max_*, angle of largest ring from optical axis; *n*, number of mires.

**Figure S2.**
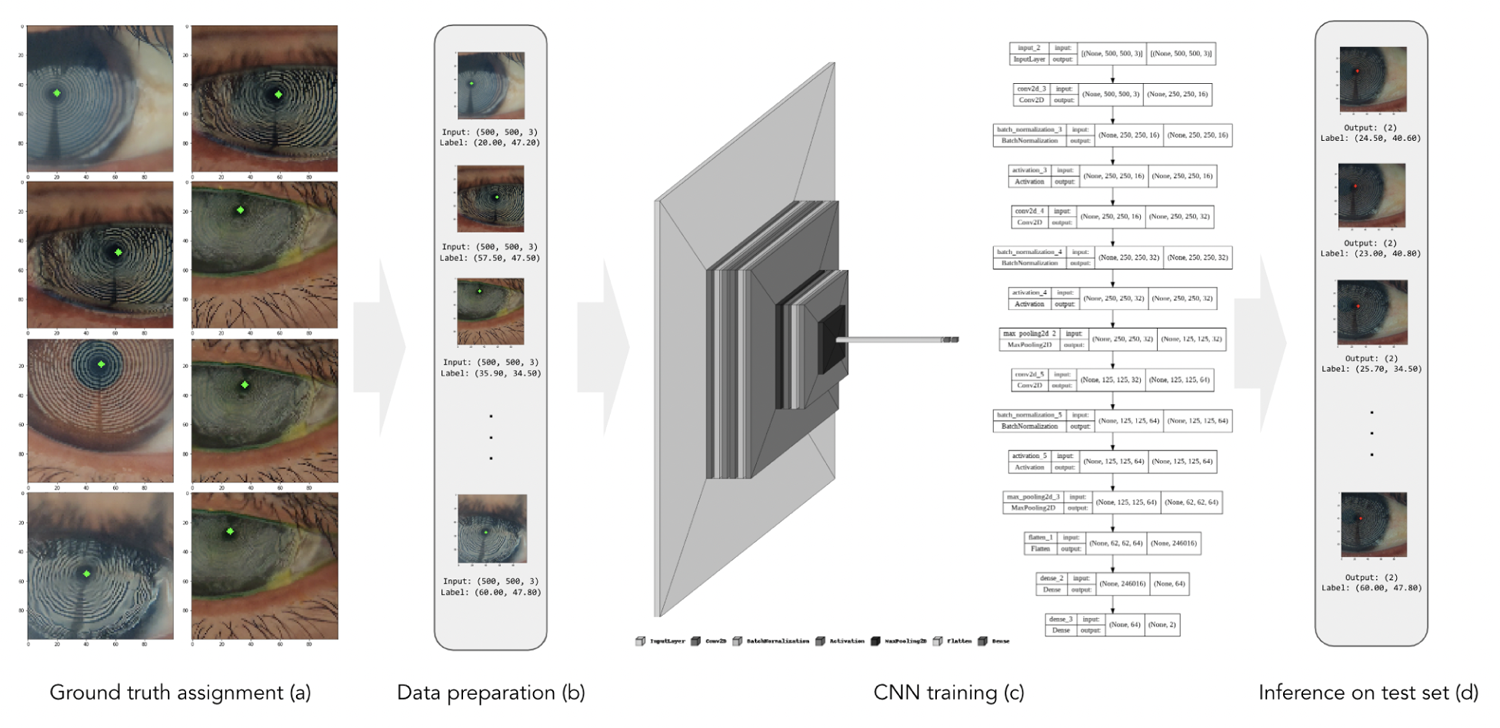
Training paradigm and architecture for center-finding regression model. (a) Ground truth assignment of mire center. (b) Preparation of data with images and ground truth. (c) Architecture of regression CNN. (d) Example results of center prediction on test set.

**Figure S3.**
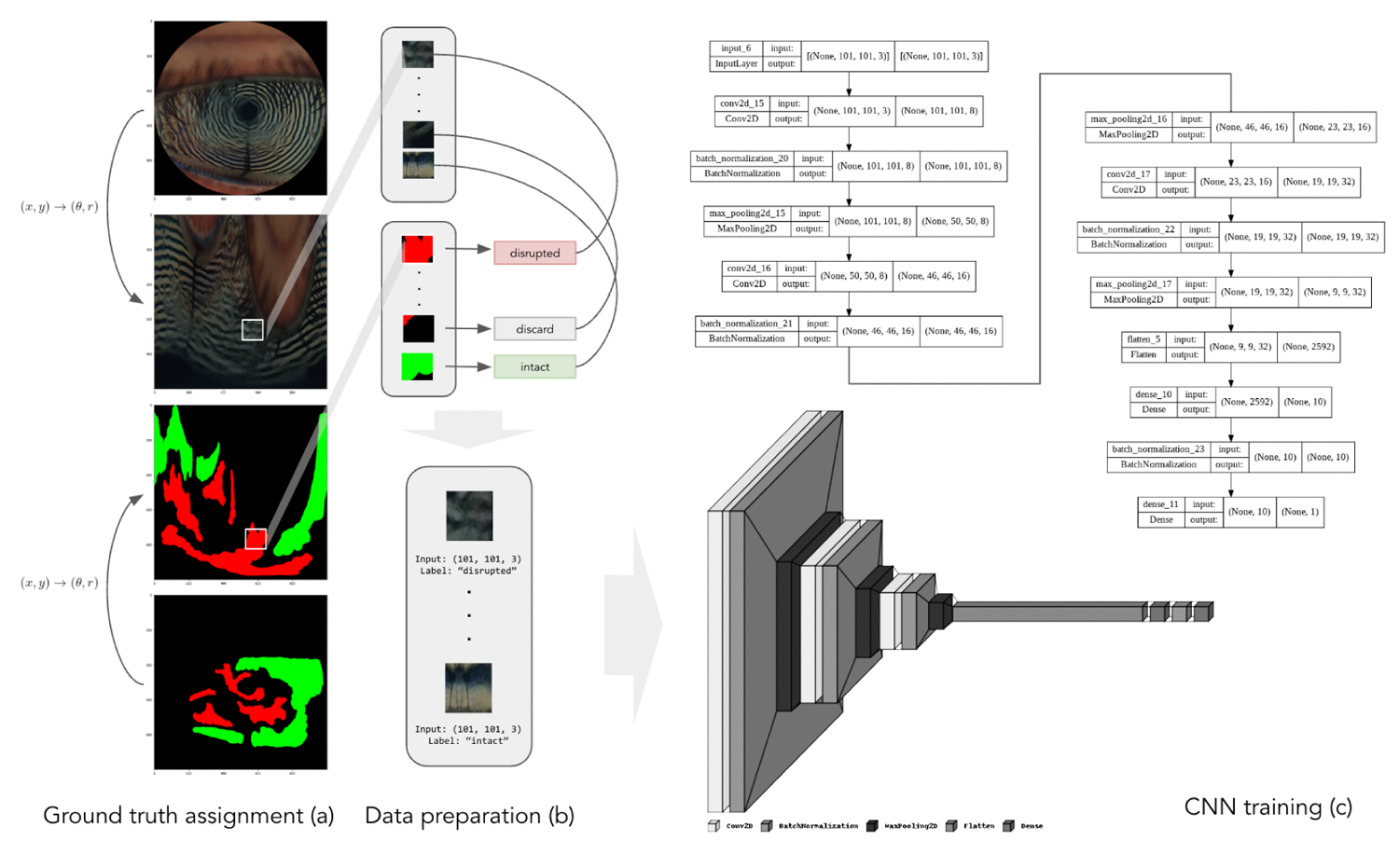
Training paradigm and architecture for patch-based classification model. (a) Ground truth assignment to patches. (b) Preparation of data with image patches and ground truth. (c) Architecture of binary classifier CNN.

## Notes

### Competing Interest Statement

The authors have declared no competing interest.

### Author Declarations

The Institutional Review Board of the L.V. Prasad Eye Institute gave ethical approval for this work, and informed consent was obtained from all subjects.

